# Molecular epidemiology of *Blastocystis* in urban and periurban human populations in Arequipa, Peru

**DOI:** 10.1101/2020.06.17.20134064

**Authors:** Kasandra Ascuña-Durand, Renzo S. Salazar-Sánchez, Ricardo Cartillo-Neyra, Jorge Ballón-Echegaray

**Author notes:** Corresponding Author: Kasandra Ascuña Durand, Laboratorio de Microbiología Molecular, Facultad de Medicina, Universidad Nacional de San Agustín, Av. Daniel Alcides Carrión S/N, Arequipa, 04000, Peru.

## Abstract

*Blastocystis* is one of the most common parasites found in the gut of humans and other hosts. It has a wide genetic diversity distributed around the world, predominating subtypes 1, 2 and 3 in South America countries. Nonspecific and inconsistent symptoms associated with this parasite made it controversial. The aim of this study was to estimate the prevalence of subtypes and determinate the epidemiological conditions associated with them. A total of 116 *Blastocystis* positive stool samples were processed by conventional PCR with *Blastocystis*-specific primers for subtypes 1 to 3. We identified subtype 1 (10.3%), subtype 2 (7.6%), subtype 3 (25.0%) and mixed infections (8.6%). We did not identify these *Blastocystis* subtypes in 48.7% of stool samples, assuming the presence of other subtypes in the zone. Any association was found between gastrointestinal symptoms and single subtype infections neither with mixed subtypes coinfections. However, our results suggest an association of *Blastocystis* subtype 2 and Irritable Bowel Syndrome (IBS, p=0.039). Besides, there was not an association between *Blastocystis* subtypes 1, 2 and 3 nor-mixed infections with epidemiological variables such as gender, age, presence of animals or vectors, places of food consumption, type of water consumption and water supply.

## Introduction

*Blastocystis* is a protozoa found in the gut of humans and animals (1,2), but it remains unclear whether it is pathogenic (3). Some researchers report an association between *Blastocystis* and gastrointestinal symptoms and diseases (4,5) whereas other investigations fail to prove any correlation between both (6,7) or suggest *Blastocystis* could be consider as normal or beneficial for the microbiota (8,9).

*Blastocystis* is genetically diverse with 17 known subtypes (2) distributed throughout the world (10). Subtypes 1–9 are found in both humans and animals, subtype 9 is only found in humans (11) and subtypes 10–17 are found in domestics and wild animals (12). Subtypes 1, 2 and 3 are the most common subtypes associated to gastrointestinal symptoms (13,14) and the most prevalent in South America (15), while subtype 4 is mainly found Europe and Asia (14). Furthermore, *Blastocystis* transmission is generally associated with poor access to healthcare and unsanitary living conditions (11,16).

Although *Blastocystis* has been studied widely over the last decade, there have been few studies focused on this parasite in South America (17,18) with only one record from Peru related to *Blastocystis* subtypes (15). *Blastocystis* has been reported in some Peruvian cities (19–21) and Arequipa has the highest prevalence of 48.29 to 81.9% in elementary school children and low income localities (22,23). Our study aims to identify subtypes 1, 2 and 3 of *Blastocystis* in urban and periurban human populations in Arequipa city and to estimate their prevalence and their association with gastrointestinal symptoms and sanitary living conditions.

## MATERIAL AND METHODS

### Ethical statement

The protocol of this study was reviewed and approved by the Institutional Review Boards of the Universidad Peruana Cayetano Heredia, reference number 18006. Written informed consent for all participants was provide prior the study. Written assent and their parents’ informed consent was provided by minors under age of 18 years old prior the study participation.

### Study site

Arequipa city is the second most populous in Peru, with approximately a million inhabitants, located in the country’s southern highlands at 2400 meters above sea level (24). The untidy growth of the city in last decades stablished two kinds of well-defined locations: periurban and urban localities, where periurban localities grew around the city with some limitation to access to basic services such as water, sanitation, health and education whereas urban areas have comprehensive access to these services. Thus, the socioeconomic status of participants from periurban areas is generally lower than urban participants (24,25).

### Study population

Stools samples came from *Blastocystis*-positive adults and children, the recruitment method is described in a previous report (26). All participants completed epidemiological surveys to assess clinical and sanitary living conditions. We obtained stools from 119 *Blastocystis*-positive participants with and without gastrointestinal symptoms.

### Detection of Blastocystis

All fresh samples were analyzed by rapid concentration with saline solution and direct observation method by standard microscopy and confirmed with blue methylene-stained stool smear under 1000X (26). *Blastocystis* positive samples were aliquot in cryovials and stored at −80°C.

### DNA extraction and PCR amplification

The DNA was extracted from fresh stool using the Norgen Stool DNA Isolation Kit (Norgen, Biotek Corporation) and stored at −20°C. PCR was performed with the 2X PCR Taq Plus MasterMix (Applied Biological Materials Inc ABM) using *Blastocystis*-specific primers F:5′-GAAGGACTCTCTGACGATGA-3’/R:5′-GTCCAAATGAAAGGCAGC-3’ (351 bp) for subtype 1 (4), F:5′-CATGAGTAAAGTCCCGTWGGGA-3’/R:5′-CCCTTTTACAGTTCATTCGCCTA-3’ (1500 bp) for subtype 2 (27), and F:5′-TAGGATTTGGTGTTTGGAGA-3’/R:5′-TTAGAAGTGAAGGAGATGGAAG-3’ (526 bp) for subtype 3 (4) with the PCR setting parameters described in figure 1. Electrophoresis was performed by adding 8 µl of the PCR products to a 1.7% agarose gel and staining with GelRed® Nucleic Acid Gel Stain, 10,000X in Water (Biotium Inc.) for 30 min at 100 V.

**Figure 1.**
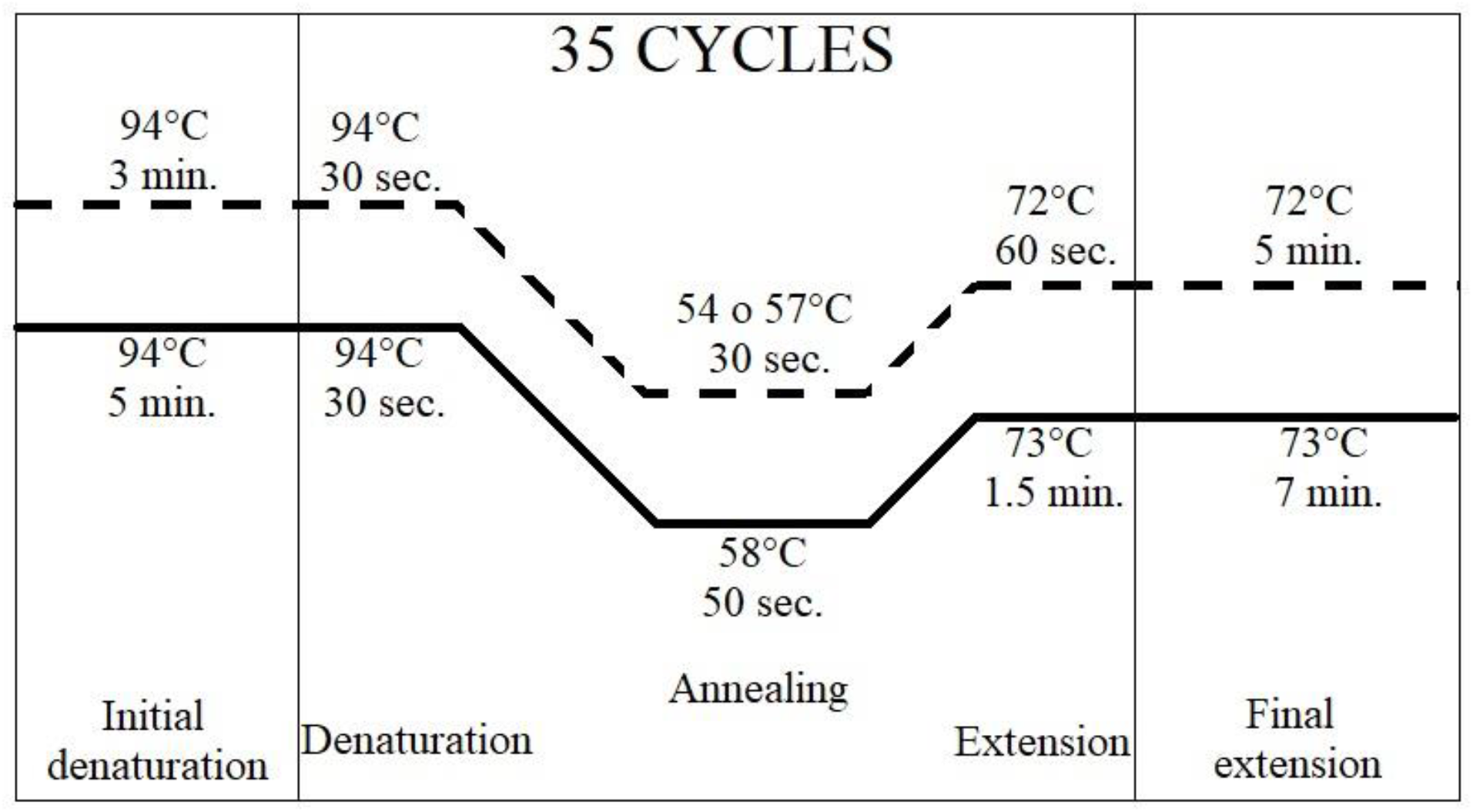
PCR setting: Cycling parameters for subtype 1 and 3 are represented in bold dashed line, for subtype 2 is in bold solid line.

### Statistical analysis

The association of *Blastocystis* subtypes with gastrointestinal symptoms and sanitary living conditions were analyzed with Chi-square and Fisher’s exact test. *Blastocystis* subtypes and patient’s age were analyzed by Kruskal-Wallis test. For the epidemiological analysis, we restricted the data to those participants from urban and periurban communities (8 samples from rural areas were excluded) and selected variables that could explain the possible source of *Blastocystis* subtypes. All analyses were conducted using R 3.6.2 (28).

## Results

We identified 119 *Blastocystis* positive individuals by standard microscopy including 50 co-infected with other parasites and commensals organisms. We were able to obtain DNA from 116 stool samples. Descriptive analysis of demographic characteristics from the selected participants shows ranged in age from 2 to 82 years old, with similar gender proportions (50.4% Female and 49.6% Male).

The PCR analysis identified samples with simple and mixed subtype infections (43.1% for simple infection and 8.6% for mixed infection). *Blastocystis* subtype 3 was the most prevalent in the study sample, followed by subtypes 1 and 2. The most common mixed subtypes identified was subtypes 1 and 3 (Table 1). We could not identify the subtypes 1, 2 or 3 of *Blastocystis* in almost half of the samples (48.7%) (Table 1). A higher frequency of subtype 3 was observed in urban and periurban localities. Subtype 2 was less frequent in periurban localities and subtype 1 in urban localities, besides only subtype 1 was identified in participants from rural localities (figure 2).

**Table 1:** Frequency and prevalence of *Blastocystis* subtypes and type of infection identified in the tested samples.

| Type of *Blastocystis*<br>infection | *Blastocystis*<br>subtype | Number of<br>participants | Prevalence<br>% |
| --- | --- | --- | --- |
| Simple infection | ST1 | 12 | 10.2 |
|  | ST2 | 9 | 7.7 |
|  | ST3 | 29 | 24.8 |
|  | Unknown | 57 | 48.7 |
| Mixed infection | ST1,3 | 9 | 7.7 |
|  | ST1,2,3 | 1 | 0.9 |

**Figure 2.**
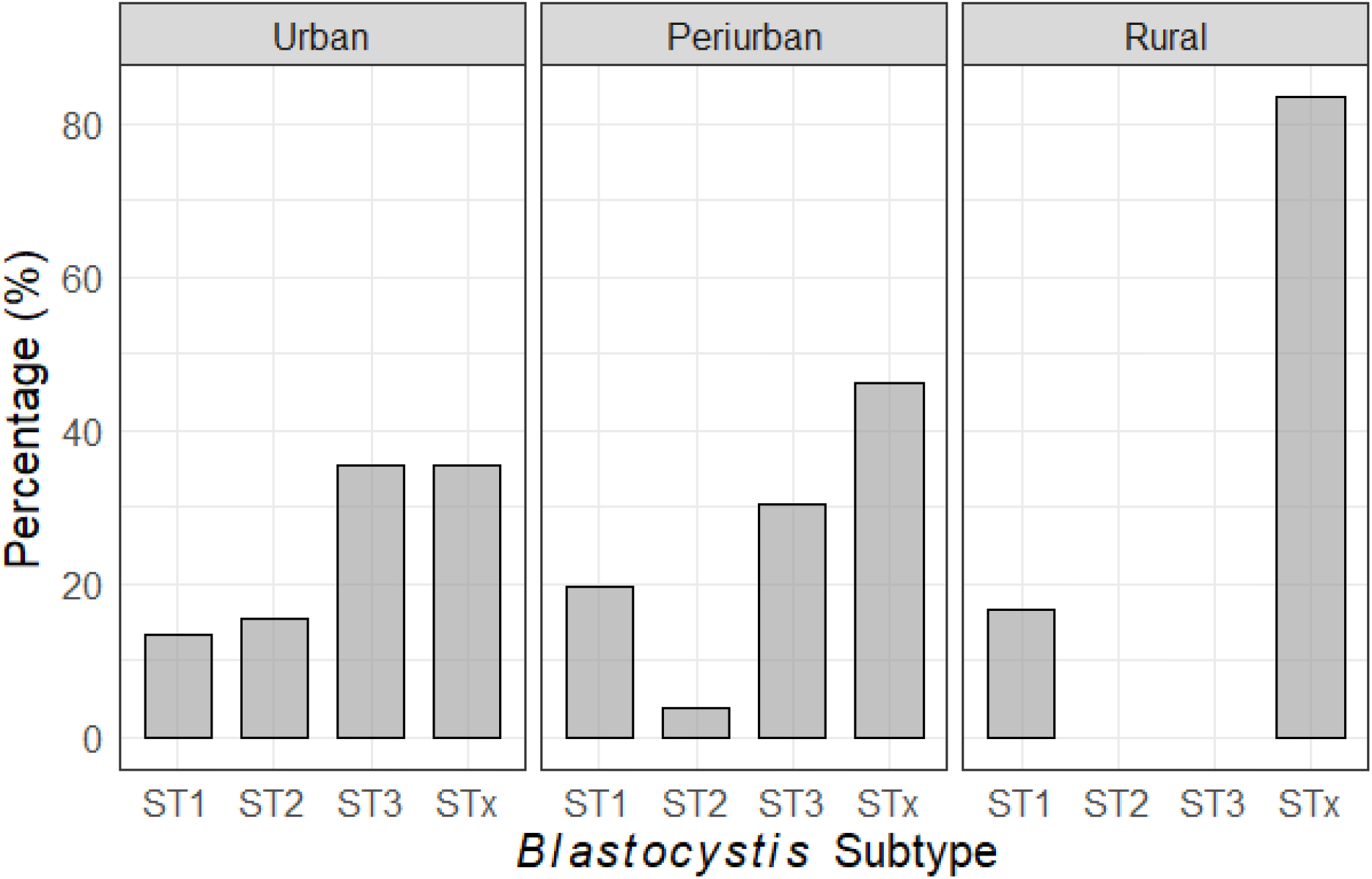
*Blastocystis* subtype distribution per kind of location. STx correspond to unknown *Blastocystis* subtype.

The descriptive analysis also shows the *Blastocystis* subtypes composition per home (n = 78 homes), where the most common composition is between subtypes 1 and 3 with 10.3% (8/78 homes), another important result was that 20 homes present only one subtype (between subtypes 1, 2 or 3) been the most prevalent subtype 3 in 15.4% of homes (12/78). A high proportion of homes had an unknown subtype (42.3%, 33/78) (figure 3 A). Additionally, we observed the presence of the 3 subtypes (1, 2 and 3) in one home in only one participant (figure 3 B), and the presence of the 3 subtypes studied and unknown subtype(s) in one home but in 4 different participants (figure 3 C).

**Figure 3.**
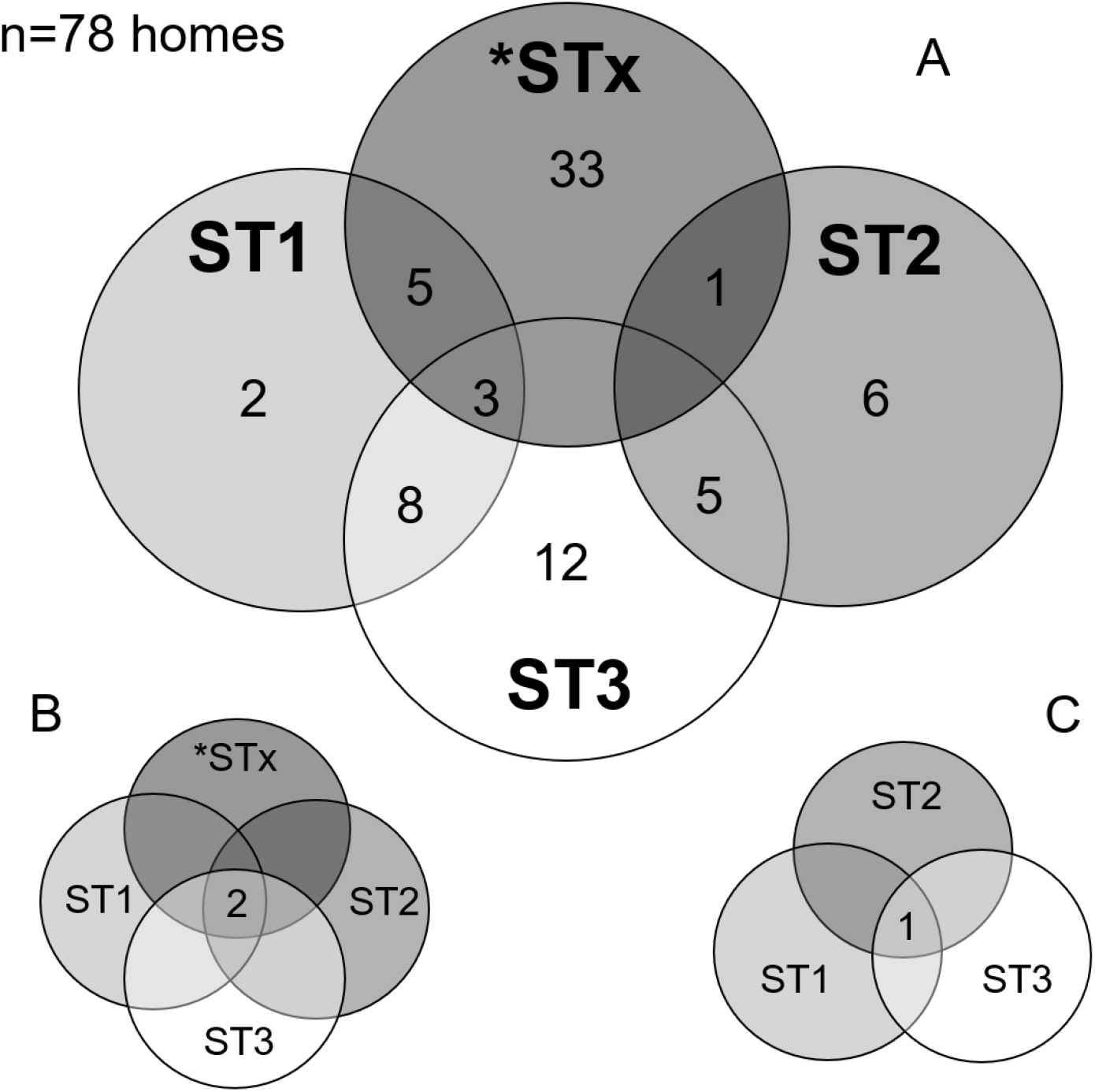
Distribution of *Blastocystis* subtypes per home. *STx: unknown subtype. A. Homes with a single, double and triple interaction between subtype 1, 2, 3 and unknown. B. Triple interaction between subtypes 1, 2 and 3. C. Quadruple interaction between subtypes 1, 2, 3 and unknown.

We also observed coinfection of *Blastocystis* subtypes with other intestinal parasites. Coinfection of *Blastocystis* subtype 3 and *Entamoeba coli* was the most frequent (57.1%), followed by *Chilomastix mesnilli* (28.6%), *Iodamoeba bütchlii* (21.4%), *Entomoeba histolytica* (14.3%) and *Giardia lamblia* (7.1%). Coinfections of *Blastocystis* subtype 1 and other parasites were found in the following frequencies: *E. coli* and *C. mesnilli* = 57.1%, *G. lamblia* = 28.8%, *I. bütchlii, E. histolytica* and *Endolimax nana* = 14.3% We did not find any coinfections associated with *Blastocystis* subtype 2.

Individuals who participated in the study lives in different districts in Arequipa city from urban and periurban areas (figure 4). Furthermore, the spatial subtype distribution shows the presence of *Blastocystis* subtype 3 in more districts than other subtypes, as the most prevalent subtype. In districts like Cerro Colorado, Cercado, M. Melgar and Characato it was the only one *Blastocystis* subtype identified. *Blastocystis* subtype 2 was present in J. Hunter and JLByR districts as the unique subtype. *Blastocystis* subtypes different than 1, 2 or 3 (unknown subtype) were present in all districts across the city. Other districts have a mix of subtypes and. Tiabaya district, located in southwestern area of the city, shows the presence of *Blastocystis* subtypes 1, 2 and 3, including unknown subtypes. The map also shows a concentration of unknown subtypes in district of Paucarpata, northeast of the city.

**Figure 4:**
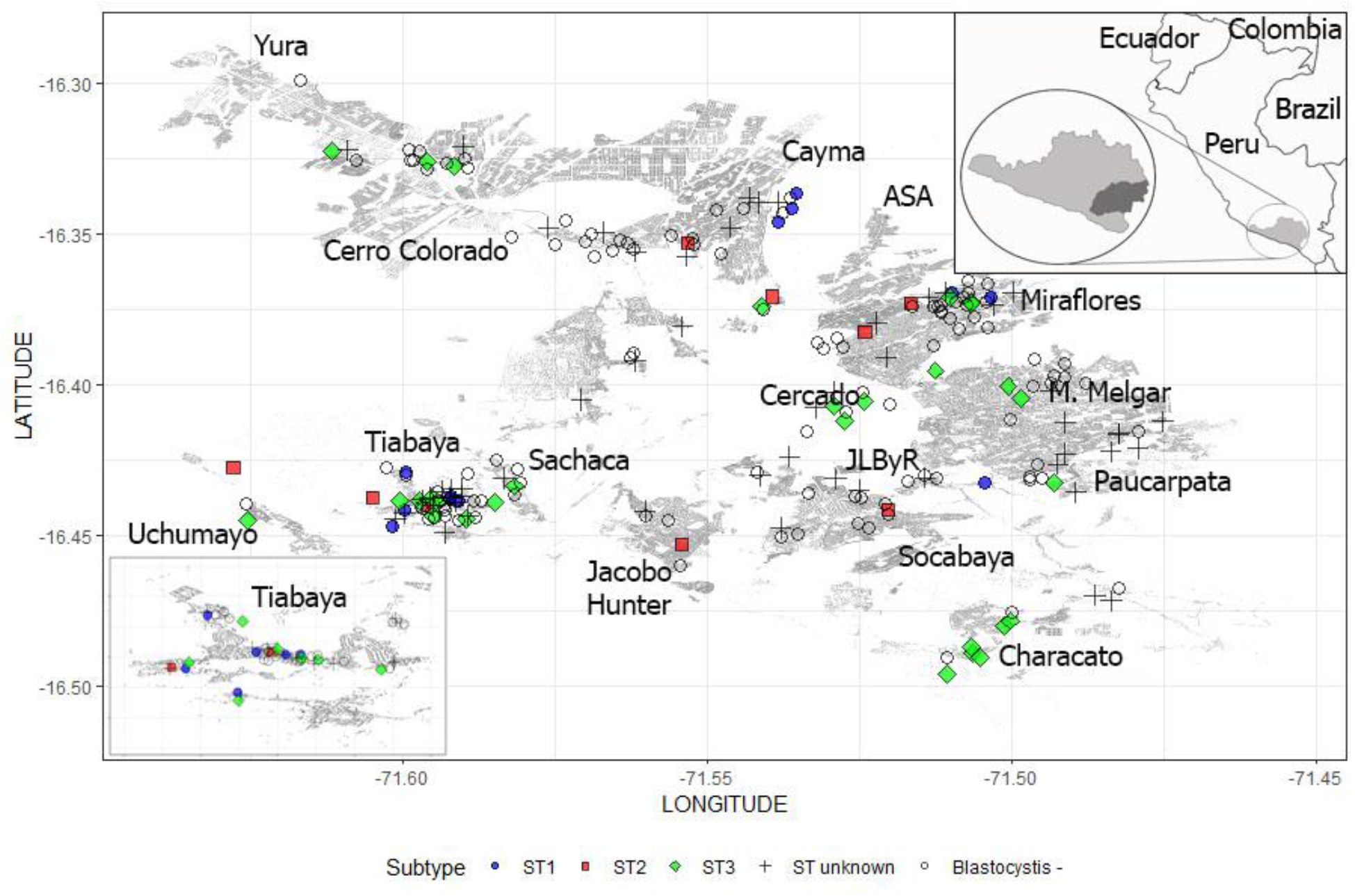
Spatial distribution of *Blastocystis* subtypes considering all the district in the metropolis with urban and periurban localities in Arequipa.

In order to exclude cross intestinal symptoms and disorders caused by other parasites, the statistical analysis of clinical manifestations was performed with *Blastocystis* positive participants and negative coinfection. Fisher exact test showed no association between single subtypes infections and the individual gastrointestinal symptoms. However, it showed a statistically significant association between *Blastocystis* subtype 2 and IBS (p 177 = 0.039). Figure 5 shows the frequency of gastrointestinal symptoms by single and mixed subtype.

**Figure 5.**
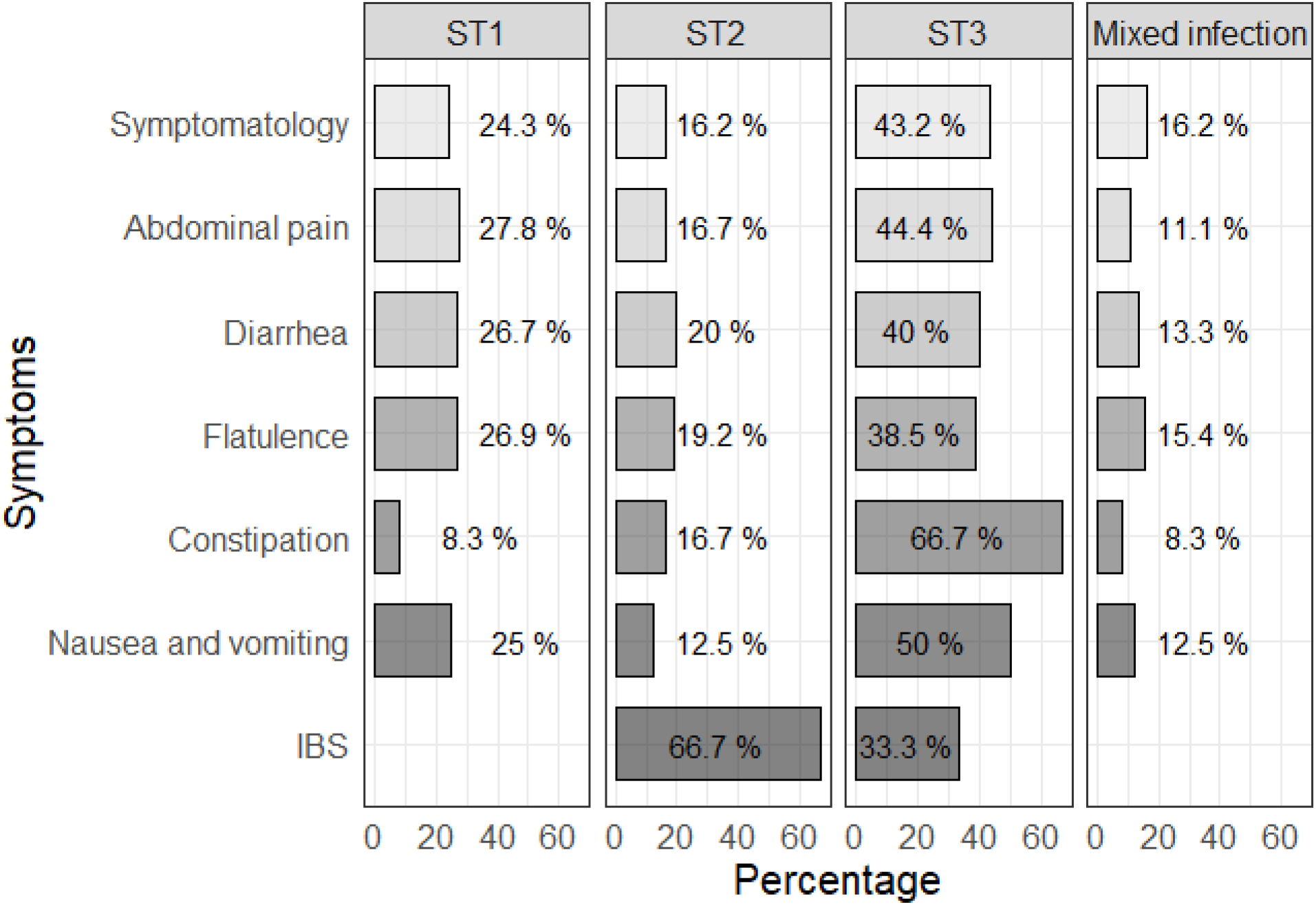
Symptomatology percentage according to *Blastocystis* subtypes 1, 2, 3 and mixed.

We included in the analysis the portion of the sample with a *Blastocystis* subtype identified, considering the mixed *Blastocystis* subtypes (n = 10) as an additional sub-group to compare. We applied a Kruskall-Wallis test to evaluate the strength in association between age of participants and subtypes in the *Blastocystis* infection, but we couldn’t find a statistical association (p.value = 0.993). We did not find a statistically significant association between single and mixed *Blastocystis* subtypes with variables from sanitary living conditions chosen for analysis (Table 2).

**Table 2:** Relationship between *Blastocystis* subtypes and sanitary living conditions.

| Variable | *Blastocystis* subtype |  |  |  | p value |
| --- | --- | --- | --- | --- | --- |
|  | ST1<br>n=21 | ST2<br>n=10 | ST3<br>n=39 | Mix infection<br>n=10 |  |
| Gender |  |  |  |  |  |
| *Female* | 16 (76.2%) | 5 (50.0%) | 21 (53.8%) | 7 (70.0%) | 0.304<sup>†</sup> |
| Age |  |  |  |  |  |
| *mean (SD)* | 32.5 (22.7) | 40.1 (23.1) | 37.3 (23.3) | 31 (26) | 0.995<sup>&</sup> |
| *median (IQR)* | 33 (11 - 49) | 46 (18 - 57) | 33 (14 - 54) | 16 (10 - 50) |  |
| Location |  |  |  |  |  |
| *Urban* | 6 (28.6%) | 7 (70.0%) | 16 (41.0%) | 5 (50.0%) | 0.178<sup>†</sup> |
| *Periurban* | 15 (71.4%) | 3 (30.0%) | 23 (59.0%) | 5 (50.0%) |  |
| Water supply |  |  |  |  |  |
| *tap water* | 20 (95.2%) | 10 (100.0%) | 36 (92.3%) | 10 (100.0%) | 1<sup>†</sup> |
| *not tap water* | 1 (4.8%) | 0 - | 3 (7.7%) | 0 - |  |
| Final feces disposal |  |  |  |  |  |
| *Piped sewer system* | 20 (95.2%) | 10 (100.0%) | 35 (89.7%) | 10 (100.0%) | 0.761<sup>†</sup> |
| *Latrine* | 1 (4.8%) | 0 - | 4 (10.3%) | 0 - |  |
| Presence of animals | 15 (71.4%) | 9 (90.0%) | 32 (82.1%) | 7 (70.0%) | 0.554<sup>†</sup> |
| Presence of dog | 12 (57.1%) | 8 (80.0%) | 26 (66.7%) | 6 (60.0%) | 0.631<sup>†</sup> |
| Presence of cat | 5 (23.8%) | 2 (20.0%) | 12 (30.8%) | 2 (20.0%) | 0.874<sup>†</sup> |
| Presence of guinea pig | 3 (14.3%) | 1 (10.0%) | 10 (25.6%) | 2 (20.0%) | 0.725<sup>†</sup> |
| Presence of rabbit | 5 (23.8%) | 2 (20.0%) | 9 (23.1%) | 2 (20.0%) | 1<sup>†</sup> |
| Presence of poultry | 5 (23.8%) | 2 (20.0%) | 14 (35.9%) | 2 (20.0%) | 0.672<sup>†</sup> |
| Presence of flies | 15 (71.4%) | 6 (60.0%) | 29 (74.4%) | 7 (70.0%) | 0.831<sup>†</sup> |
| Presence of cockroach | 5 (23.8%) | 1 (10.0%) | 9 (23.1%) | 2 (20.0%) | 0.911<sup>†</sup> |
| Presence of rodents | 3 (14.3%) | 2 (20.0%) | 7 (17.9%) | 1 (10.0%) | 1<sup>†</sup> |
| Place of food consumption |  |  |  |  |  |
| *House* | 17 (81.0%) | 7 (70.0%) | 32 (82.1%) | 8 (80.0%) | 0.377<sup>†</sup> |
| *Restaurant* | 2 (9.5%) | 1 (10.0%) | 1 (2.6%) | 1 (10.0%) |  |
| *House and restaurant* | 0 - | 2 (20.0%) | 4 (10.3%) | 0 - |  |
| *House and kiosk* | 2 (9.5%) | 0 - | 2 (5.1%) | 1 (10.0%) |  |
| Kind of water consumption |  |  |  |  |  |
| *Boiled water* | 18 (85.7%) | 8 (80.0%) | 32 (82.1%) | 9 (90.0%) | 0.737<sup>†</sup> |
| *Tap water* | 1 (4.8%) | 0 - | 0 - | 0 - |  |
| *Both* | 2 (9.5%) | 2 (20.0%) | 7 (17.9%) | 1 (10.0%) |  |

† Fisher & Kruskal-Wallis

## Discussion

This study is the first large-scale study at molecular level carried out in Peru to determinate the presence and distribution of *Blastocystis* subtypes and its relationship with clinical and epidemiological aspects in urban and periurban communities of Arequipa, Peru. We found *Blastocystis* subtypes 1, 2 and 3, where *Blastocystis* subtype 3 was the most prevalent followed by subtype 1 and 2. Similar distribution and prevalence have been reported in South America (10,15,29) and some places around the world (4,30,31). However, other studies from Brazil, and Lao Democratic Republic shows subtype 1 is the most prevalent (32,33), while in Europe the most prevalent subtypes are 3 and 4 (34,35).

The only previous molecular study of *Blastocystis* in Peru identified the presence of subtypes 3 and an unknown subtype (15) in 13 participants. In our study we used a much larger sample size and identified subtypes 1, 2, 3 and unknown subtypes. Mixed-double infections between *Blastocystis* subtype 1, 2 and 3 with different proportions and combinations have been reported in Brazil, Turkey and Iran (4,30,32). In our result we report 8.6% mixed-double subtypes infections corresponding to subtypes 1 and 3. We also report a mixed-triple subtypes infection in only one sample for *Blastocystis* subtypes 1, 2 and 3.

We also report a high number of samples with unknown subtypes that was not expected according to the previously report from Peru (15). However, we could infer that exist another subtypes present in Arequipa human population, being possible due to local migration, touristic activities (31), or even zoonotic transmission due to farming activities (10). The unknown subtypes in our study could match with other subtypes reported in surrounding countries (15) that also can be supported by migration to Arequipa city.

Respecting to migration, it has been widely studied as one of the main factor that explain the urbanization of Chagas disease, a parasitic and endemic disease from rural areas in Arequipa (36,37), where constant migration since 1970’s from rural to urban environments created a new environment with rural customs as raising domestic animals in periurban communities around the city (38). Additionally, the lack of healthy sanitary conditions could also establish proper conditions to facilitate infection and transmission of enteroparasites. These reasons also could explain the distribution of subtypes into the city suggesting that geographical features, sociocultural activities and customs combined with unappropriated sanitary conditions are not a barrier that limit this parasite and subtypes to rural or urban environments. In contrast with the reported in Turkey, where exist a well-defined distribution of subtypes 5, 6 and 7 in rural and subtypes 1, 2 and 3 in urban areas (39).

Since the pathogenicity of *Blastocystis* was suggest in 1986 (40), the controversy has not stopped due to many research indicate the association of this parasite to symptoms and diseases and others do not. In that way, some studies suggest that some subtypes such as 1 and 3 are responsible of some gastrointestinal symptoms like abdominal pain, diarrhea, vomiting, flatulence (4,5), which is not found in our study. We can highlight that in symptomatic individuals the presence of subtype 2 shows an association with IBS; whereas other study shows the same *Blastocystis* subtypes in IBS individuals (41,42). However, we could not support this association with the individual symptomatology showed by participants in our study due only four participants showed this condition syndrome. By the other hand, it was reported that *Blastocystis* subtype 3 could have a carcinogenic influence or facilitate the proliferation and growth of existing cancer cells (43,44).

According to demographic variables, the scientific literature reports statistical differences between men and women mainly for the economic activities (18). Contrary to these results, we have not found statistical association between *Blastocystis* subtypes and sex, in spite of many adult woman are dedicated to welfare home in the study sample and considering this activity as risk of infection due to food contamination (45,46), lack of sanitation (18) and raising animals in periurban communities from Arequipa (36). By the other hand, age is another important demographic factor of exposition and play a key role in intestinal infections as referred in other studies (13,39,47). In this study median age in single-subtype infections has values over 30 years old while mixed infection is more frequent under 20 and subtype 3 has a normal distribution across ages (Figure 4). Regardless of these distribution, the outcome do not show a statistical association with *Blastocystis* subtypes, as was obtained in Malaysia where subtype 2 was significantly associated to ages under or equal 15 years old (48).

Coinfection or polyparasitism has an impact on morbidity and nutrition, causing increase in susceptibility and intensity of infections and overreacting of immune response (49). We found a low percentage of coinfection with non-pathogenic and pathogenic enteroparasites as *E. coli, Ch. Mesnilli, E. histolytica* and *G. lamblia*. Other studies have reported coinfection between *Blastocystis* and *Dientamoeba fragilis* or *G. duodenalis* (29)(50). We explored coinfections in our study under the scope that some *Blastocystis* subtypes are pathogens (51) and considering that coinfection can produce an increase the intensity of infections. Nevertheless, our results on clinical manifestations and coinfection could support the hypothesis that *Blastocystis* is a beneficial microorganism and part of the healthy microbiome (52) and not a pathogen as lately research has begun to showed associating *Blastocystis* with a healthy microbiome (8,9), a rich diet in vegetables (53) and a better immune status (7).

Some studies found a positive correlation between *Blastocystis* and bacterial richness which suggests that *Blastocystis* is most common in people with healthy bacterial flora and negative correlated to Bacteroides that is observed more prevalent in unhealthy people being able to use *Blastocystis* as to determine the health of the individual rather than the bacterial composition (8,9,54).

The exploration of sanitary variables had the aim to identify the *Blastocystis* subtype provenance and to determinate possible transmission routes and sources. The overall available literature has limited references about these associations, only subtype 3 is associated to not boiled water drinking behavior (48). Other variables as food consumption and raising animals has no association to subtypes 1 and 3 (55).

We could not find any association between epidemiological variables and subtypes 1, 2 or 3 of *Blastocystis*; similar results was reported to drinking water, hand and food washing behavior (56). However in rural places, socioeconomic position, food and water combined to raising animals is the reason of specific subtypes distribution in rural communities (39). General prevalence studies on intestinal parasite infections has associated the presence of *Blastocystis* infection to untreated water consumption, final feces disposal and inadequate hands washing habits (21,45).

Almost half of the positive samples in our study had an indeterminate subtype, which is an important limitation. We assume that this outcome is related likely to primers sensitivity. Other important limitation was not to consider in the study the presence of subtypes reported in lower proportions for south America, expecting similar results as previous reported to Peru. We suggest a deeper review in large population with other molecular techniques like NGS to elucidate the indeterminate subtypes in this study. We also suggest to explore the likely association between subtype two and three with IBS in murine model to discard this pseudo association.

Despite these limitations, the data obtained herein are the first on prevalence of Blastocystis and distribution of subtypes in Arequipa and will contribute to a better understanding of the molecular epidemiological role and context of this parasite. Likewise, our findings state the basis for future research and explore new hypothesis about distribution and clinical manifestations of Blastocystis subtypes in our region.

## Conclusions

We report the presence of Blastocystis subtypes 1, 2 and 3 were detected in Arequipa communities, as widely reported in South America and other countries around the world. Infection with we suggest an association between *Blastocystis* subtype 2 and was associated with IBS. The statistical analysis shows that No significant association was found between sanitary living conditions and specific *Blastocystis* subtypes.

## Data Availability

The data can be requested by email to the fist author.

## Acknowledgments

We thanks to Zoonotic Diseases Research Laboratory from Universidad Peruana Cayetano Heredia (ZDRL-UPCH) for permission in the use of georeferenced data of Arequipa city to build the map of *Blastocystis* subtypes distribution, to Drs. Irmia Paz, Victor Vásquez, Elí Martinez and Mr. Cirilo Neyra (UNSA) for their helpful contribution to this project.

